# Transmission interval estimates suggest pre-symptomatic spread of COVID-19

**DOI:** 10.1101/2020.03.03.20029983

**Authors:** Lauren C. Tindale, Michelle Coombe, Jessica E. Stockdale, Emma S. Garlock, Wing Yin Venus Lau, Manu Saraswat, Yen-Hsiang Brian Lee, Louxin Zhang, Dongxuan Chen, Jacco Wallinga, Caroline Colijn

## Abstract

**Background:** As the COVID-19 epidemic is spreading, incoming data allows us to quantify values of key variables that determine the transmission and the effort required to control the epidemic. We determine the incubation period and serial interval distribution for transmission clusters in Singapore and in Tianjin. We infer the basic reproduction number and identify the extent of pre-symptomatic transmission.

**Methods:** We collected outbreak information from Singapore and Tianjin, China, reported from Jan.19-Feb.26 and Jan.21-Feb.27, respectively. We estimated incubation periods and serial intervals in both populations.

**Results:** The mean incubation period was 7.1 (6.13, 8.25) days for Singapore and 9 (7.92, 10.2) days for Tianjin. Both datasets had shorter incubation periods for earlier-occurring cases. The mean serial interval was 4.56 (2.69, 6.42) days for Singapore and 4.22 (3.43, 5.01) for Tianjin. We inferred that early in the outbreaks, infection was transmitted on average 2.55 and 2.89 days before symptom onset (Singapore, Tianjin). The estimated basic reproduction number for Singapore was 1.97 (1.45, 2.48) secondary cases per infective; for Tianjin it was 1.87 (1.65, 2.09) secondary cases per infective.

**Conclusions:** Estimated serial intervals are shorter than incubation periods in both Singapore and Tianjin, suggesting that pre-symptomatic transmission is occurring. Shorter serial intervals lead to lower estimates of R0, which suggest that half of all secondary infections should be prevented to control spread.

## 1 Introduction

The novel Coronavirus Disease, COVID-19, was first identified in Wuhan, Hubei Province, China in December 2019 and has since spread across the globe. Many of the initial cases were traced to the Huanan Seafood Wholesale Market [1], however the first patient (symptom onset December 1, 2020) did not have exposure to the market [2]. Wuhan health authorities closed the market on January 1, 2020, first isolated SARS-CoV-2 from a patient on January 7, 2020 [3], and publicly released the genome sequence on January 10, 2020 [4]. This prompt release of genomic data has allowed public health authorities globally to implement screening and disease preparedness procedures and to develop diagnostic workflows to confirm SARS-CoV-2 infections [5, 6].

The serial interval can be used to estimate the basic reproduction number (R0), which is used to determine the amount of transmission that needs to be stopped in order to contain an outbreak. The serial interval and incubation period distributions together can be used to identify the extent of pre-symptomatic transmission. Early COVID-19 estimates borrowed parameters from SARS [7, 8, 9], but more recent estimates have been made using information from early clusters of COVID-19 cases. Depending on the population used, estimates for incubation periods have ranged from 3.6-6.4 days and serial intervals have ranged from 4.0-7.5 days [1, 10, 11, 12, 13]; however, it is crucial that the estimates of incubation period and serial interval are based on the same outbreak, and are compared to those obtained from outbreaks in other populations. New distinct outbreak clusters are ideal for understanding how COVID-19 can spread through a population with no prior exposure to the virus.

The first Singapore COVID-19 case was confirmed as an individual who had travelled to Singapore from Wuhan. Many of the initial cases were imported from Wuhan, with later cases being caused by local transmission. Singaporean officials have been working to identify potential contacts of confirmed cases; close contacts were monitored and quarantined for 14 days from their last exposure to the patient, and other low risk contacts were put under active surveillance and contacted daily to monitor their health status.

Tianjin is a city in the northeast of China with a population of over 15 million. COVID-19 cases were traced to a department store, where numerous customers and sales associates were likely infected. Additional customers who had potential contact were asked to come forward through state news and social media, as well as asked if they had visited the department store at various checkpoints in the city. All individuals identified as having visited the store in late January were quarantined and sections of the Baodi District where the store is located were sealed and put under security patrol.

We screened publicly available data to identify datasets for two COVID-19 clusters that can be used to estimate transmission dynamics within a relatively closed system, where immediate public health responses were implemented, contacts were identified and quarantined, and key infection dates were tracked and published. We use these clusters to estimate both incubation period and serial interval for COVID-19, in both datasets, and discuss the implications of our findings for R0 estimates and pre-symptomatic transmission.

## 2 Methods

### 2.1 Data

All datasets and R code are available on GitHub (github.com/carolinecolijn/ClustersCOVID19). Singapore data was obtained from the Ministry of Health Singapore [14] online press releases. The Singapore dataset comprised 93 confirmed cases from the date of the initial case on January 19, 2020 until February 26, 2020. Tianjin data was obtained from the Tianjin Health Commission [15] online press releases. The Tianjin dataset comprises 135 cases confirmed from January 21 to February 22, 2020. The symptom onsets were available on the official website for all but a few patients who had not had symptoms before being diagnosed at a quarantine center. Both datasets contained information on exposure times, contacts among cases, time of symptom onset and more (see Supplementary Information for column descriptions and data processing).

### 2.2 Statistical analysis

For both Singapore and Tianjin, the daily incidence of hospitalization and mortality was plotted with the cumulative number of cases confirmed and discharged. The daily incidence rate was also visualized by date of symptom onset per probable source of infection. For the symptom onset plots, any cases that did not have information on date of onset of symptoms were removed. Cases were then grouped based on information provided in the “presumed reason” column for Singapore dataset and by the “Infection source” column in the Tianjin dataset. In the Tianjin dataset, there were a small number of cases (n = 10) that could be classified into two possible infection source groups (e.g. from Wuhan and has a close relationship with another known case). These cases were assigned their infection source groups based on the following hierarchy of possible sources: (highest priority) Known relationship > Wuhan origin > Other China travel > Location unclear travel > Mall (for shoppers, workers, or individuals living near to the Baodi mall outbreak) > Unknown (lowest priority). All cases in the Singapore dataset were categorized into an infection source group without conflict. The group designations were not used in the statistical estimates.

Incubation period estimates were based on data for the earliest and latest possible exposure times, and on the reported times of symptom onset. It is impossible to confirm the exact times of exposure, so we use interval censoring (R package icenReg [16]) to make parametric estimates of the incubation period distribution. Where the earliest exposure is not specified in the “start source” column, we noted that the case must have been exposed some time since the beginning of the outbreak (Dec 01, 2019). Where the last possible exposure is not given in “end source”, we noted that exposure must have occurred before symptom onset. In the Singapore dataset, some cases had a travel history or contact with a known location or presumed source of the virus and this defined a most recent time of exposure. For these we proceed as in Tianjin. Other individuals had estimated exposure times based on the symptom times for their presumed infector. For these, we define an exposure window using the symptoms of their presumed infector ±4 days. Having defined exposure windows, we proceed with interval censoring as in the Tianjin data. In both datasets we stratified the data according to whether symptom onset occurred early or late and estimated incubation periods separately.

Serial intervals were estimated with the expectation-maximization approach described in Vink et al [17]. Briefly, this approach assigns the case with earliest symptom onset in the cluster as a “putative index” (PI) status, and uses the time difference between symptom onset of subsequent cases in the cluster and the putative index as “index case to case” (ICC) intervals for putative index cases in small clusters. The ICC intervals are the time differences between the symptom onset in the putative index (PI) and the others, so the collection of times *t*_*j*_ − *t*_*pi*_ for each *j* in the cluster. These intervals are not samples of the serial interval distribution, because it need not be the case that the PI infected the others. Vink et al [17] used a mixture model in which ICC intervals *t*_*j*_ − *t*_*pi*_ can arise in four ways: (1) an outside case infects PI and *j*; (2) PI infects *j*; (3) PI infects an unknown who infects *j* and (4) PI infects unknown 1 who infects unknown 2 who infects *j*. Accordingly, if the serial interval *x* ∼ 𝒩 (*µ, σ*^2^), the density for the ICC intervals is

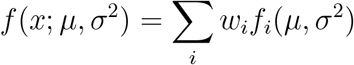

where *w*_*i*_ are weights of the *i*′*th* component density and *f*_*i*_ are the component densities for the *i*′*th* transmission route.See [17] for more details. Expectation-maximization is used to determine *µ* and *σ*.

For each dataset we create a graph whose nodes are individuals and whose edges are reported direct contacts between individuals. We use the connected components of the graph to define the small clusters. We also explored using earliest end exposure time, in case the first symptomatic case was not the index case for the cluster. A few clusters are large enough that the four models in the mixture are likely insufficient to cover the transmission possibilities, so we restrict the analysis to only the first 3, 4, 5 or 6 cases per cluster.

The mean time difference between transmitting infection by an infector and symptom onset is (assuming independence) then the difference between the mean serial interval and the mean incubation time. We calculate the reproduction number as in [18]: *R* = exp *rµ* − 1/2*r*^2^*σ*^2^ with *r* the exponential growth rate, *µ* the mean serial interval and *σ* the standard deviation of the serial interval. To obtain confidence intervals for *R* we resample *µ* and *σ* from our bootstrap samples; this preserved covariation of *µ* and *σ*. Statistical analyses were performed using R [19].

## 3 Results

### 3.2 Descriptive Analyses

Figures 1 and 2 show the daily counts, putative origin of the exposure and individual time courses for the Singapore and Tianjin data. In the Singapore dataset, new hospitalization and discharge cases were documented daily from January 22 to February 26, 2020. 66.7% (62/93) of the confirmed cases recovered and were discharged from the hospital by the end of the study period (Figure 1a). The disease progression timeline of the 93 documented cases in Figure 1c indicates that symptom onset occurred 6.6 ± 4.8 (mean ± SD) days after the initial presumed viral exposure and hospitalization occurred 5.9 ± 5.1 days after symptom onset. The mean length of hospital stay was 13.3 ± 6.0 days before individuals recovered and were discharged.

**Figure 1:**
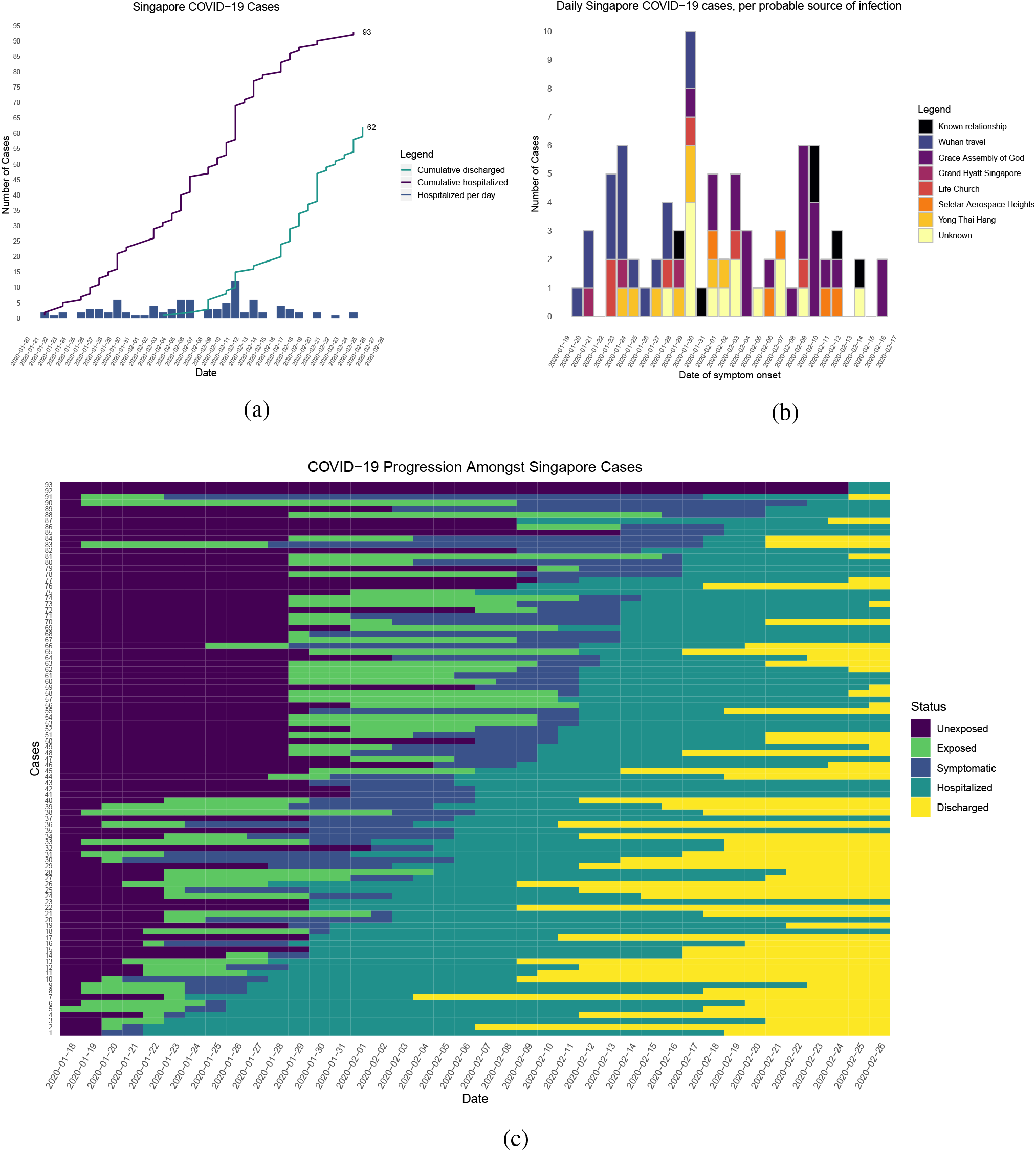
Singapore COVID-19 cases. (a) Daily hospitalized cases and cumulative hospitalized and discharged cases. (b) Incidence curve with probable source of infection (c) Disease progression timeline, including dates at which each case is unexposed, exposed, symptomatic, hospitalized, and discharged. Not all cases go through each status as a result of missing dates for some cases.

**Figure 2:**
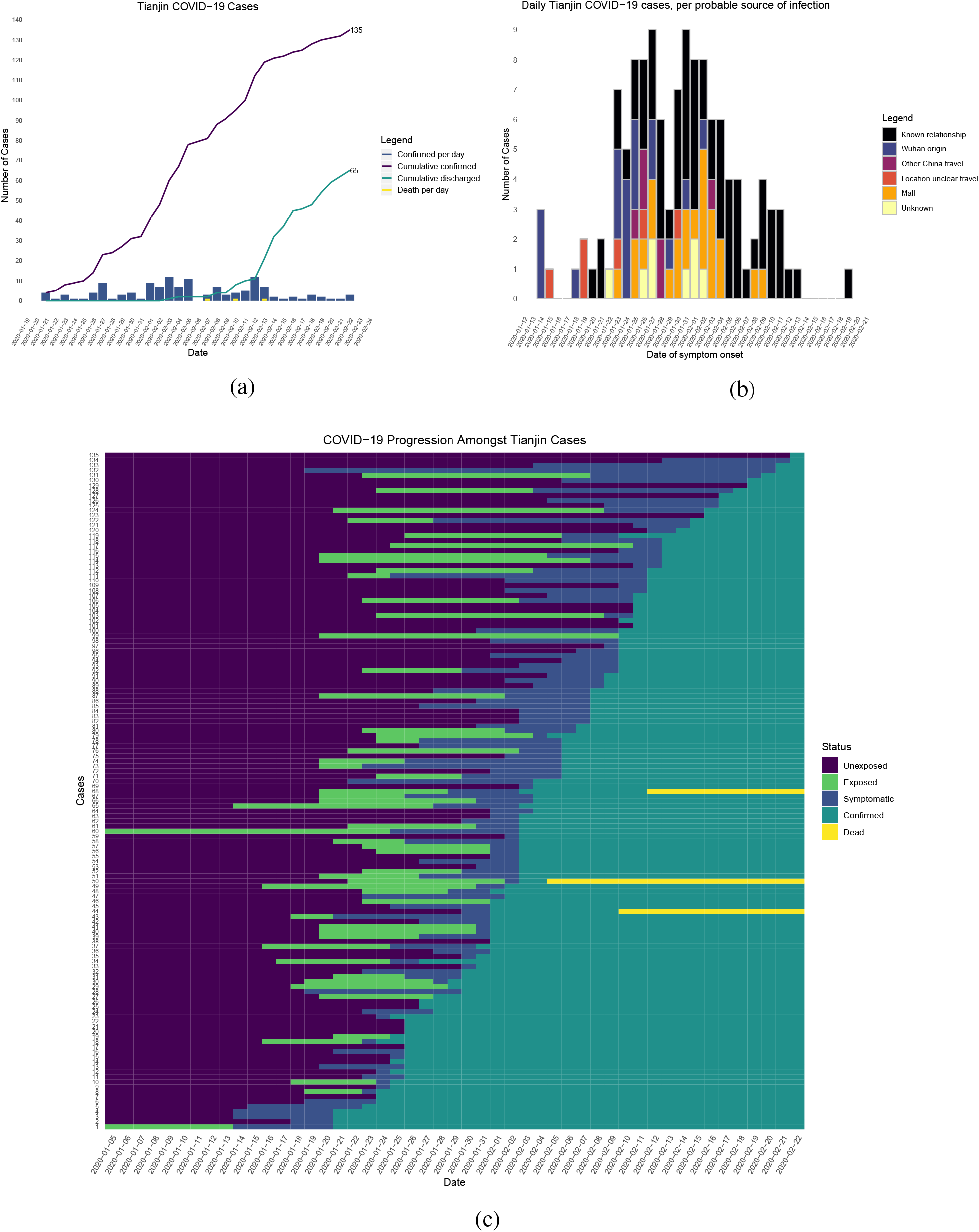
Tianjin COVID-19 cases. (a) Daily and cumulative confirmed cases, cumulative discharges and daily death cases. (b) Incidence curve with probable source of infection (c) Disease progression timeline; not all cases go through each status as a result of missing dates for some cases

In the Tianjin dataset, new confirmed cases were documented daily from January 21 to February 22, 2020. 48.1% (65/135) recovered and 2.2% (3/135) had died by the end of the study period (Figure 2a). The timeline of the 135 cases is shown in Figure 2c. Symptom onset occurred 5.4 ± 4.5 (mean ± SD) days after the latest presumed viral exposure. Cases were confirmed 5.2 ± 4.2 days after symptom onset. The duration of hospital stay of the Tianjin cases is unknown as the discharge date of each case was not available. In both datasets, daily counts decline over time, which is likely a combination of delays to symptom onset and between symptom onset and reporting, combined with the effects of strong social distancing and contact tracing.

### 3.2 Incubation period

In the Singapore dataset, we find that the median incubation period is 6.55 days with the Weibull distribution; shape 1.88 (95%CI 1.47, 2.39); and scale 7.97 (6.8, 9.3). The mean incubation period is 7.1 (95%CI 6.1, 8.3) days. In Tianjin we find a median 8.62 days; shape 2.25 (1.79, 2.83); scale 10.15 (8.91, 11.56). The mean is 9 (7.92, 10.2) days. We also fitted gamma and log normal distributions; see Tables 1 and S1. These are consistent with previous estimates.

**Table 1:**
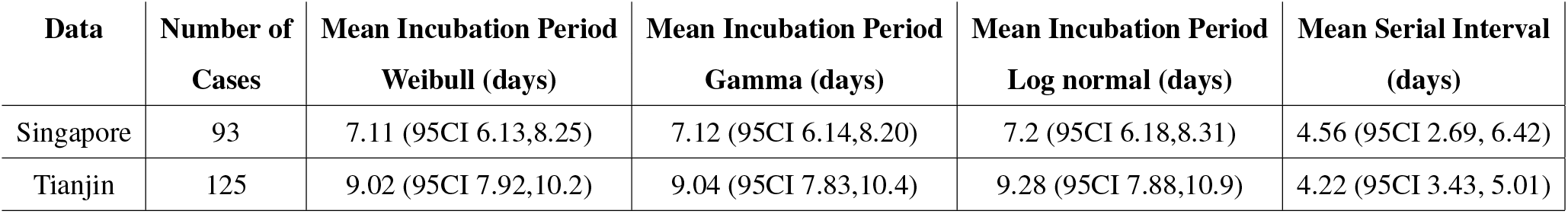
Mean incubation period and mean serial interval estimated for COVID-19 using Singapore and Tianjin datasets. CIs are based on the standard deviation of bootstrap estimates of the mean serial interval.

| **Data** | **Number of Cases** | **Mean Incubation Period Weibull (days)** | **Mean Incubation Period Gamma (days)** | **Mean Incubation Period Log normal (days)** | **Mean Serial Interval (days)** |
| --- | --- | --- | --- | --- | --- |
| Singapore | 93 | 7.11 (95CI 6.13,8.25) | 7.12 (95CI 6.14,8.20) | 7.2 (95CI 6.18,8.31) | 4.56 (95CI 2.69, 6.42) |
| Tianjin | 125 | 9.02 (95CI 7.92,10.2) | 9.04 (95CI 7.83,10.4) | 9.28 (95CI 7.88,10.9) | 4.22 (95CI 3.43, 5.01) |

However, in Singapore they based on a combination of cases for whom last possible exposure is given by travel, and later cases (for whom the presumed infector was used). In Tianjin, social distancing measures took place. We find that the estimated incubation period is different, particularly in Tianjin, for cases with symptom onset prior to January 31. The estimated median incubation period for pre-Jan 31 cases in Tianjin is 6.9 days; the *q* = (0.025, 0.975) quantiles are (2, 12.7) days. In contrast, post-Jan 31 the median is 12.4 days with *q* = (0.025, 0.975) quantiles (5.4, 19) days. The means are 7 (6.1, 8.0) days for early cases and 12.4 (10.8, 14.2) days for later cases. Social distancing seems unlikely to change the natural course of infection, but these results might be explained if exposure occurred during group quarantine or otherwise later than the last time individuals thought they could have been exposed. Pre-symptomatic transmission would enable this.

In Singapore we find the same effect, though less pronounced. The estimated median incubation time is 5.46, with (0.025, 0.975) quantiles of (1.34, 11.1) days for early cases and 7.27 days (quantiles (1.31, 17.3)) days for late-arising cases. The means are 5.71 (4.55, 7.06) days for early cases and 7.86 (6.57, 9.38) days for later cases. Fits of gamma and log-normal distributions are similar; see Table S2. In Singapore, this difference could be explained by the fact that presumed infectors’ symptom times are used to define exposure windows, but infection may be indirect; however, changes in perception of exposure times and pre-symptomatic transmission could result in missing intermediate transmission events and hence lengthened incubation period estimates.

### 3.3 Serial interval, pre-symptomatic transmission and R0

Table 1 shows our ICC estimates of the mean and standard deviation for the serial intervals, with comparison to other analyses and assumptions in Table S4. We estimate the mean serial interval to be 4.22 (3.43, 5.01) days with standard deviation 0.40 for Tianjin and 4.56 (2.69, 6.42) days (sd 0.95) for Singapore, using the first 4 cases in each cluster. Du et al [20] obtain a similar estimate for the serial interval (3.96 days with 95%CI: 3.53-4.39) but with standard deviation 4.75 days, based on 468 cases in 18 provinces.

Our serial intervals are notably shorter than our incubation period estimates, suggesting that there is pre-symptomatic transmission, with infection occurring on average 2.89 and 2.55 days before symptom onset of the infector (Tianjin, Singapore). Because the incubation period is different for early- and late-occurring cases in our data, on average transmission for early-occurring cases is 2.79 and 1.2 days before symptom onset (Tianjin, Singapore) and 8.2, 3.3 days before (Tianjin, Singapore) for late-occurring cases. The fact that serial intervals are shorter than incubation periods is robust in our sensitivity analysis (Table S3). These estimates are strengthened by the fact that we have estimated both incubation period and serial interval in the same population and by the fact that we obtain the same result in two distinct datasets. In both sets of estimates, samples of the incubation period minus serial interval are negative with probability 0.8 or higher (Tianjin) and 0.7 or higher (Singapore), suggesting that a substantial portion of transmission may occur before symptom onset (see Supplementary Information and Figure S2), consistent with the clinical observations reported by Rothe et al. [21] and Bai et al. [22].

Shorter serial intervals yield lower reproduction number estimates. For example, when the epidemic grows at a rate of 0.15 (doubling time of 6.6 days; [23] scenario 1), the reproduction number for Tianjin is *R* = 1.87 (1.65, 2.07) and for Singapore it is 1.97 (1.43, 2.51). In contrast, if the previous serial interval (7.5 days [23, 1]) is used, the estimate is *R* = 3.05.

## 4 Discussion

Here we use transmission clusters in two locations where cases have reported links, exposure and symptom onset times to estimate both the incubation period and serial interval of COVID-19. These are integral parameters for disease forecasting and for informing public health interventions. Serial intervals, together with R0, control the shape and distribution of the epidemic curve [24]. They influence the disease’s incidence and prevalence, how quickly an epidemic grows, and how quickly intervention methods need to be implemented by public health officials to control the disease [24, 25]. In particular, the portion of transmission events that occur before symptom onset is a central quantity for infection control [25].

In Singapore and Tianjin we estimated relatively short serial intervals. Of particular note, early estimates of R0 for COVID-19 used the SARS serial interval of 8.4 days [7, 8, 26]. Our serial interval findings from two populations mirror those of and Zhao et al [27] and Nishiura et al [13], who estimated a serial interval of 4.4 and 4.0 days. Furthermore, we estimate the serial interval to be shorter than the incubation period in both clusters, which suggests pre-symptomatic transmission. This indicates that spread of SARS-CoV-2 is likely to be difficult to stop by isolation of detected cases alone. However, shorter serial intervals also lead to lower estimates of R0, and our serial intervals support R0 values close to 2; if correct this means that half of the transmissions need to be prevented to contain outbreaks.

We stratified the incubation period analysis for Tianjin by time of symptom onset (pre- or post-Jan 31, 2020; motivated by quarantine/social distancing measures) and found that the apparent incubation period was longer for those with post-quarantine symptom onset. The reason for this is unclear, but one possible explanation is that there were (unknown, therefore unreported) exposures during the quarantine period. If people are quarantined in groups of (presumed) uninfected cases, pre-symptomatic transmission in quarantine would result in true exposure times that are more recent than reported last possible exposure times.

There are several limitations to this work. First, the times of exposure and the presumed infectors are uncertain, and the incubation period is variable. We have not incorporated uncertainty in the dates of symptom onset. We have used the mixture model approach for serial intervals to avoid assuming that the presumed infector is always the true infector, but the mixture does not capture all possible transmission configurations. Our R0 estimates are very simple and could be refined with more sophisticated modelling in combination with case count data. We have not adjusted for truncation (eg shorter serial intervals are likely to be observed first). However, the serial interval estimates are consistent between the two datasets, are robust to the parameter choices, and are consistently shorter than the estimated incubation times.

In conclusion, in both the Singapore and Tianjin COVID-19 clusters we identified both the incubation period and the serial interval. Our results suggest that there is substantial pre-symptomatic transmission, with the serial interval shorter than incubation period by 2-4 days. We find differences in estimated incubation period between early and later cases; this may be due to pre-symptomatic transmission or differences in reporting and/or in perceived exposure as the outbreak progressed, in the context of social distancing measures. Finally, our shorter serial intervals lead to an estimate of the basic reproduction number of approximately 2 in both datasets, suggesting that stopping half the transmission events may be sufficient to control outbreaks.

## Data Availability

Data are available at the link below.

https://github.com/carolinecolijn/ClustersCOVID19

## Acknowledgements

We thank the Ministry of Health Singapore [14] and the Tianjin Health Commission [15] for publishing information about cases through online press releases. We thank the participants of ‘Epi-CoronaHack’ at Simon Fraser University for their roles in curating these two datasets. We thank the public health teams and the patients whose data have been included in these analyses.

## Supplementary Appendix

### 1 Details of the Singapore and Tianjin datasets

In the Singapore dataset: “related cases” are direct known contacts between cases; “cluster links” are cases that are linked together through an identified cluster event; “presumed infected date” and “presumed reason” are the earliest known date and the reason that each case was known to likely be infected; “last poss exposure” and “symptom presumed infector” are sub-classifications of “presumed infected date,” representing either the last date that each case could have been infected – the date of arrival in Singapore for travellers from Wuhan – or the date that each case was likely infected during a local transmission event in Singapore, respectively; “cluster” is the Ministry of Health Singapore’s classification of cases into transmission cluster events.

In the Tianjin cases summary spreadsheet, the main columns are: gender, age, symptom onset, symptom type, confirmation date, severity and death date [1]; detailed information from daily reports for the first 80 patients provided travel or exposure history and contact information, from which we obtained exposure windows (start source, end source). For backup and to complete missing information for later cases we also referred to Jinyun News, Tianjin official local media: [2] who used Baodi local government reports [3]. They reported detailed activity for those confirmed cases when their corresponding epidemiological history investigation was finished.

### 2 Statistical methods

#### 2.1 Incubation period

We explored several distributions for the incubation period: Weibull, gamma and log normal. As shown in Figure 3, once fit the resulting distributions all provide very similar results. Table S1 summarizes the parameter estimates for the gamma and log normal distributions. Table S2 gives the parameters for the incubation period for early- and late-occurring cases in both datasets.

**Figure 3:**
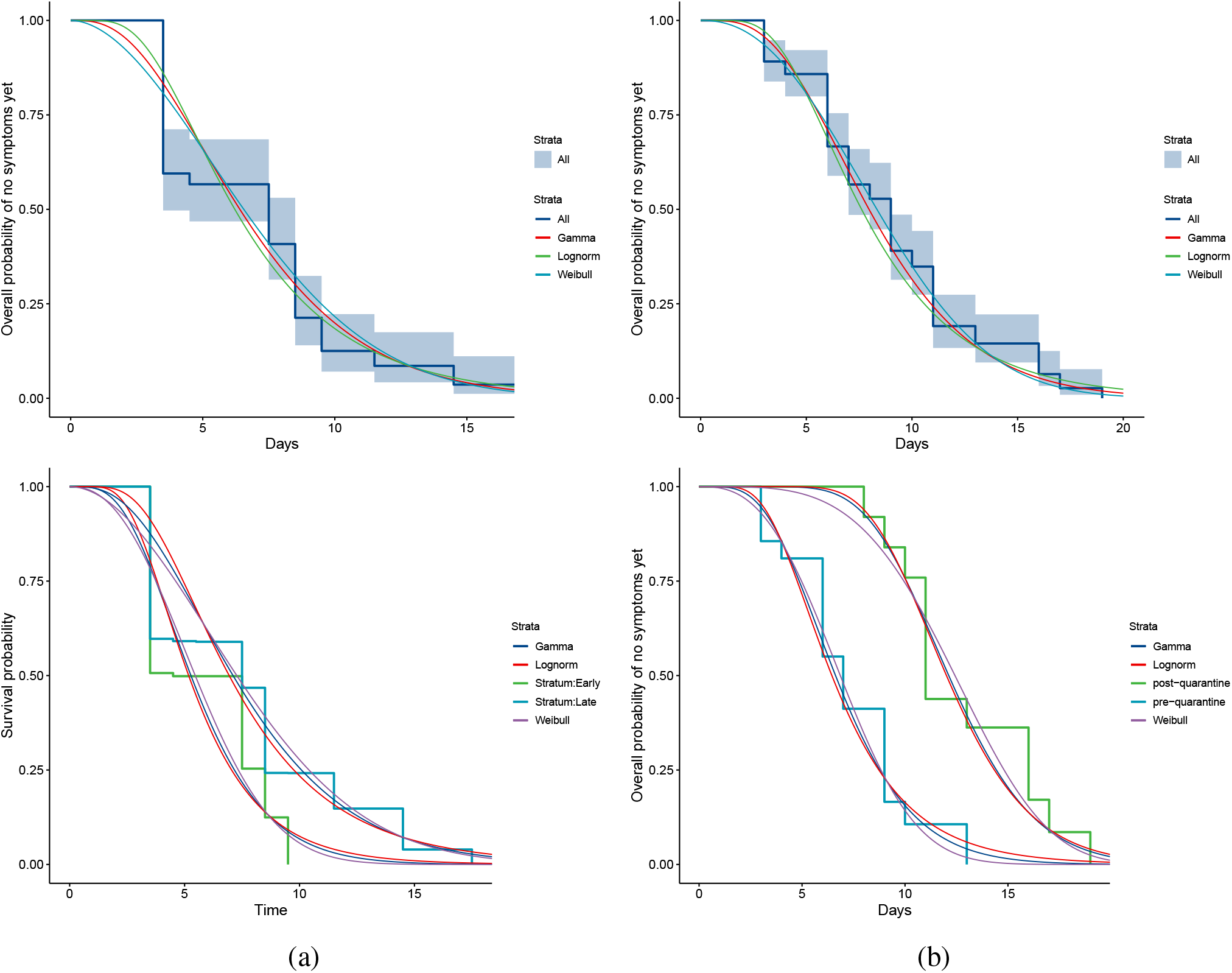
COVID-19 incubation period Kaplan-Meier curves for (a) Singapore and (b) Tianjin. Top panels show unstratified data (all cases with symptom onset given). Bottom panels show ‘early’ and ‘late’ cases, defined in Tianjin by those with symptom onset prior to January 31 vs later, and in Singapore, those with a specified last possible exposure (all prior to or on January 30).

**Figure 4:**
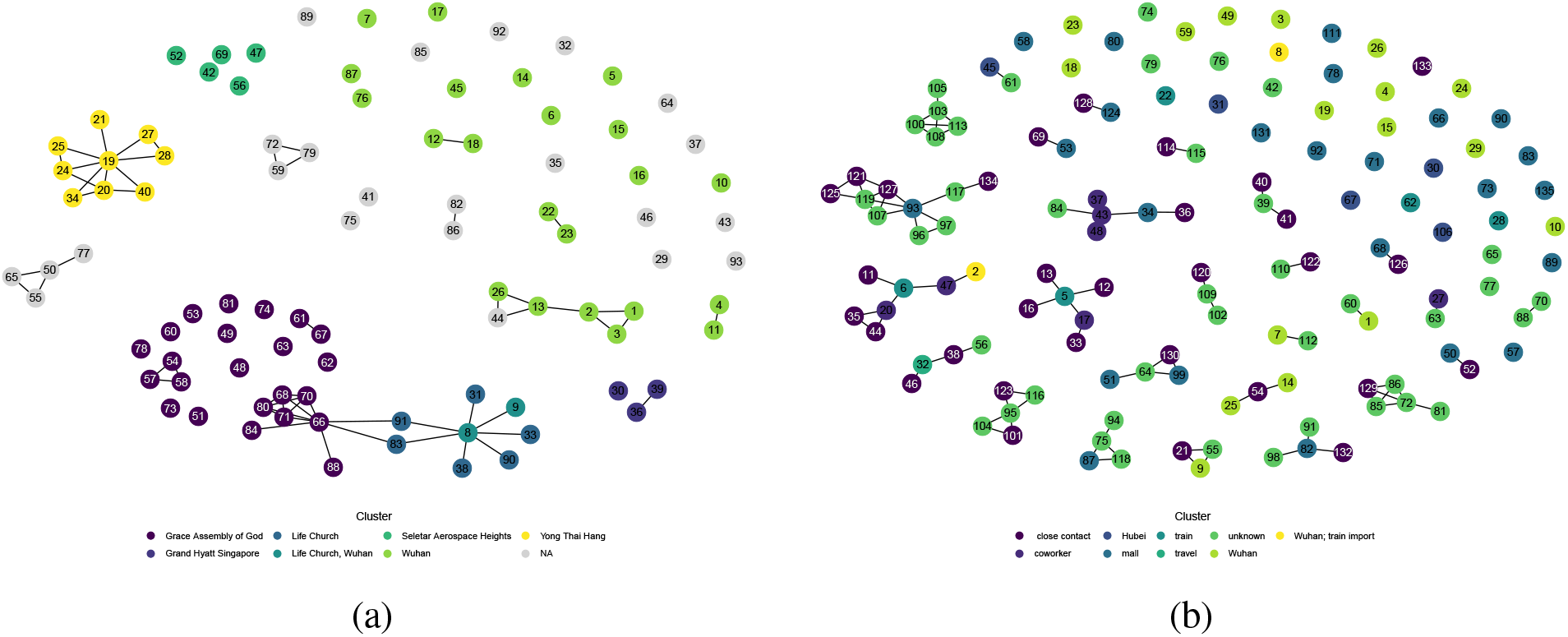
Network diagram for (a) Singapore (b) Tianjin.

#### 2.2 Serial interval

We used bootstrapping to explore the range for the point estimates of *µ* and *σ* from the mixture model. Figure S1 shows the results. The mean of the bootstrapped mean estimates is 4.26 ± 1.02 for Tianjin and 4.57 ± 0.95 days in Singapore. Bootstrap values are consistent with a serial interval that is considerably shorter than the incubation periods in both datasets. Table S3 shows the sensitivity analysis; we varied the the number of cases per cluster to include in the ICC interval data and we explored sorting the cases in the clusters according to the time of last exposure (ie the putative index status assigned to the individual with the earliest end to their exposure window, instead of the first symptomatic individual).

#### 2.3 Pre-symptomatic transmission

We sampled the estimated serial interval and incubation period distributions to estimate the probability that the incubation period minus the serial interval is negative. This could indicate the portion of transmission that occurs before symptom onset, but it makes the strong assumption that incubation period and onset of symptoms occur independently and it does not take the growth curve of the outbreak, or censoring or truncation, into account. We find that the portion of negative samples is 0.67 for early cases and 0.73 for unstratified (overall incubation period) cases in Singapore, and 0.82 (early) and 0.87 (all) cases in Tianjin. Early cases have shorter estimated incubation periods so the fractions are lower. Late cases have higher portions (0.78, 0.98) but this is due to the long incubation estimate and fixed serial interval distribution; it may be influenced by censoring and truncation. Figure S2 illustrates the distributions.

### 3 Additional published estimations

Estimates of incubation period and serial interval from other studies are shown in Table S4. Of note, the majority of studies do not estimate both incubation period and serial interval in the same population.

**Figure S1:**
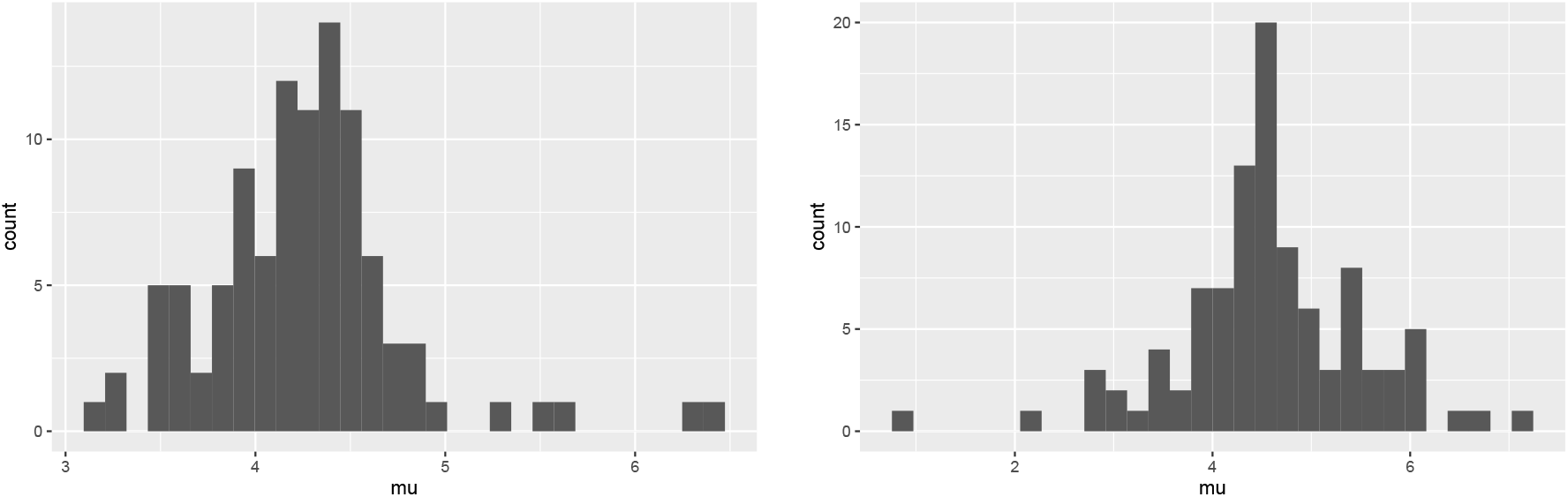
Bootstrap values of the mean serial interval for (left) Tianjin and (right) Singapore, based on 100 replicates using the first 4 cases in each cluster.

**Figure S2:**
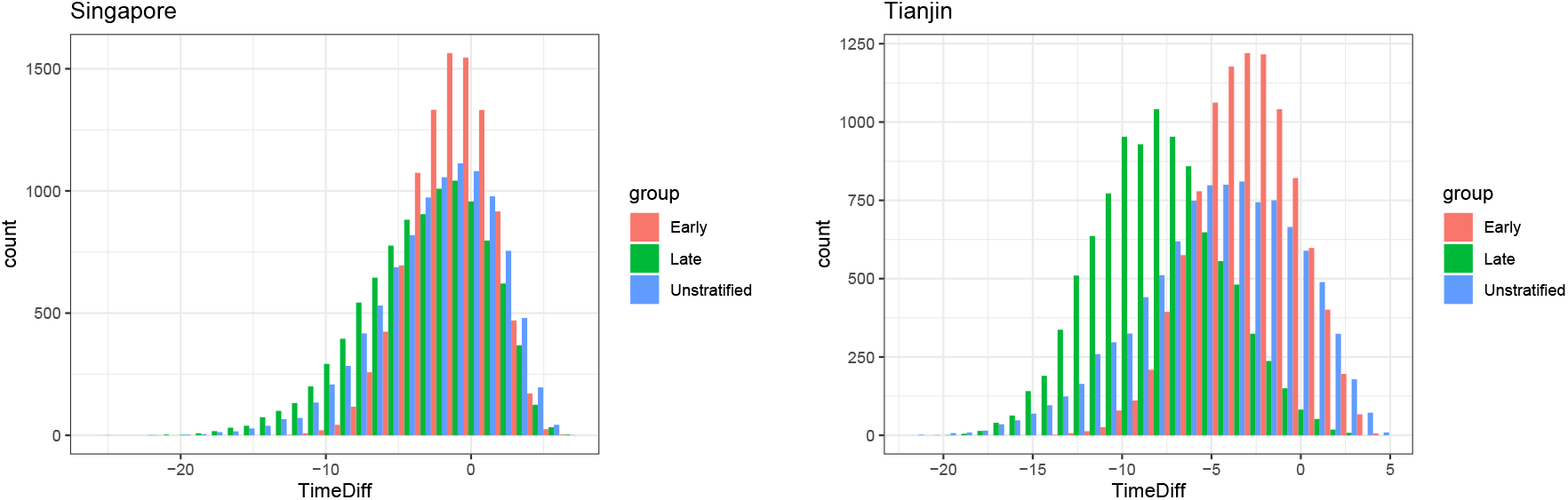
Samples of the incubation period distribution minus the serial interval distribution, assuming independence and without considering the (unknown) growth curve of the outbreaks or the extent of censoring.

**Table S1:**
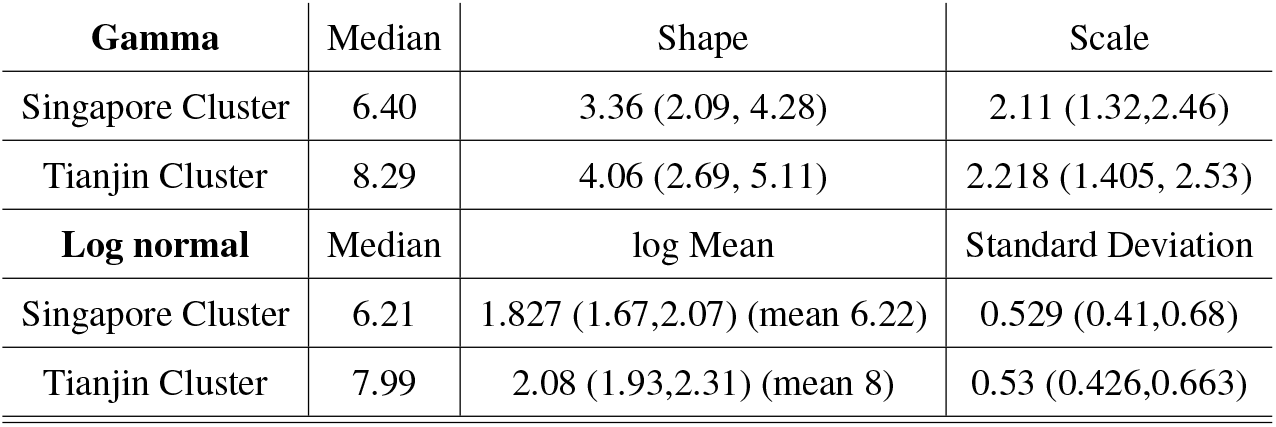
Incubation period estimates using a gamma distribution. 95% confidence intervals for the shape and scale (log mean and sd for log normal) parameters are shown in brackets.

| **Gamma** | Median | Shape | Scale |
| --- | --- | --- | --- |
| Singapore Cluster | 6.40 | 3.36 (2.09, 4.28) | 2.11 (1.32,2.46) |
| Tianjin Cluster | 8.29 | 4.06 (2.69, 5.11) | 2.218 (1.405, 2.53) |
| **Log normal** | Median | log Mean | Standard Deviation |
| Singapore Cluster | 6.21 | 1.827 (1.67,2.07) (mean 6.22) | 0.529 (0.41,0.68) |
| Tianjin Cluster | 7.99 | 2.08 (1.93,2.31) (mean 8) | 0.53 (0.426,0.663) |

**Table S2:** Incubation period estimates using stratified data

| **Tianjin** |  |  |  |
| --- | --- | --- | --- |
| Weibull | Median | Shape | Scale |
| Early | 6.857 | 2.73 (2.012, 3.725) | 7.84 (6.851, 8.972) |
| Late | 12.4 | 3.95 (2.662, 5.853) | 13.636 (11.987, 15.51) |
| Gamma | Median | Shape | Scale |
| Early | 6.65 | 5.42 (3.13,6.82) | 1.31 (0.716,1.49) |
| Late | 12.1 | 14.33 (6.61,18.03) | 0.866 (0.395,0.986) |
| Log normal | Median | log mean | standard deviation |
| Early | 6.51 | 1.87 (1.72,2.01) | 0.449 (0.345,0.585) |
| Late | 12.0 | 2.49 (2.36,2.72) | 0.263 (0.178,0.39) |
| **Singapore** |  |  |  |
| Weibull | Median | Shape | Scale |
| Early | 5.46 | 2.35 (1.5,3) | 6.38 (5.14,7.43) |
| Late | 7.27 | 1.93 (1.43,2.46) | 8.79 (7.35,10.2) |
| Gamma | Median | Shape | Scale |
| Early | 5.28 | 4.64 (1.94, 5.92) | 1.22 (0.53,1.42) |
| Late | 7.11 | 3.57 (1.96,6.52) | 2.19 (1.22,2.55) |
| Log normal | Median | log mean | standard deviation |
| Early | 5.18 | 1.644 (1.41,1.88) | 0.457 (0.286,0.729) |
| Late | 6.94 | 1.94 (1.75,2.12) | 0.504 (0.372,0.684) |

**Table S3:** Serial interval estimates

| ordering | Number cases per cluster | $\mu$ (Tianjin) | $\sigma$ (Tianjin) | $\mu$ (Singapore) | $\sigma$ (Singapore) |
| --- | --- | --- | --- | --- | --- |
| Onset | 3 | 4.03 | 1.05 | 4.36 | 1.11 |
| Onset | 4 | 4.22 | 0.978 | 4.56 | 1.16 |
| Onset | 5 | 4.32 | 1.028 | 4.81 | 1.24 |
| Onset | 6 | 4.43 | 1.071 | 5.02. | 1.28 |
| Last Exposure | 4 | 4.51 | 0.94 | 3.92 | 1.23 |
| Bootstrap | 4 | 4.23 (sd 0.402) | 0.952 (sd 0.172) | 4.57 (sd 0.95) | 1.24 (sd 0.411) |

**Table S4:** Mean incubation period and mean serial interval estimates for COVID-19 generated by other studies.

| Data | Number of Cases | Mean Incubation Period<br>(days) | Mean Serial Interval<br>(days) | Reference |
| --- | --- | --- | --- | --- |
| Wuhan first cases | 425 | 5.2 (95CI 4.1-7.0) | 7.5 (95CI 5.3-19) | Li et al. 2020 [4] |
| South Korea first cases | 24 | 3.6 | 4.6 | Ki et al. 2020 [5] |
| Travellers from Wuhan | 88 | 6.4 (95CI 5.6-7.7) | - | Backer et al. 2020 [6] |
| Diagnosis outside Wuhan<br>(excluding Wuhan residents) | 52 | 5.0 (95CI 4.2-6.0) | - | Linton et al. 2020 [7] |
| Diagnosis outside Wuhan<br>(including Wuhan residents) | 158 | 5.6 (95CI 5.0-6.3) | - | Linton et al. 2020 [7] |
| Transmission chains<br>in Hong Kong | 21 chains | - | 4.4 (95CI 2.9-6.7) | Zhao et al 2020 [8] |
| Infector-infectee pairs* | 28 pairs | - | 4.0 (95CI 3.1-4.9) | Nishiura et al 2020 [9] |

\*Note: included 3 infector-infectee pairs from this Singapore cluster. Remainder from Vietnam (4), South Korea (7), Germany (4), Taiwan (1) and China (9)

